# Trace, Quarantine, Test, Isolate and Treat: A Kerala Model of Covid-19 Response

**DOI:** 10.1101/2020.06.15.20132308

**Authors:** KM Sulaiman, T Muhammad, Rishad Muhammad A P, K Afsal

**Affiliations:** International institute for Population Sciences-MUMBAI

**Keywords:** Covid-19, Kerala, India, Testing, Tracing, Pandemic

## Abstract

Kerala reported the first three cases of coronavirus in India in late January. Kerala, one of the India’s most densely populated states, which makes its success in fighting the Covid-19 all the more commendable. Moreover, an estimated 17% of its 35 million population employed or lives elsewhere, more than 1 million tourists visit each year, and hundreds of students study abroad, including in China. All of this mobility makes the state more vulnerable to contagious outbreaks. What is the strategy behind the success story? This paper compares the situation of COVID-19 pandemic in major states and Kerala by the different phase of lockdown, and also highlights Kerala’s fight against the pandemic. We used publicly available data from https://www.covid19india.org/ and Covid-19 Daily Bulletin (Jan 31-May 31), Directorate of Health Services, Kerala (https://dashboard.kerala.gov.in/). We calculate the phase-wise period prevalence rate (PPR) and the case fatality rate (CFR) of the last phase. Compared to other major states, Kerala showed better response in preventing pandemic. The equation for the Kerala’s success has been simple, prioritized testing, widespread contact tracing, and promoting social distance. They also imposed uncompromising controls, were supported by an excellent healthcare system, government accountability, transparency, public trust, civil rights and importantly the decentralized governance and strong grass-root level institutions. The “proactive” measures taken by Kerala such as early detection of cases and extensive social support measures can be a “model for India and the world”.

## Introduction

Since the first case of COVID-19 reported at the end of January, India has reported more than 250,000 Covid-19 infections. More than 7,000 people have died due to the virus. As of 31 May, the testing positivity rate in the country was near 4%, the death rate from the pandemic was around 3% and the doubling rate of disease - or the amount of time it takes to double the Covid-19 infections was 12 days. 40% recovery rate of infected patients was reported in this period (1). Same as other countries, there are hotspots and clusters of infection in India. Nearly 80% of the active cases are in five major states - Maharashtra, Tamil Nadu, Delhi, Gujarat and Madhya Pradesh, and importantly more than 60% of the cases in five cities, including Mumbai, Delhi, Chennai and Ahmedabad, according to Government reports (2). India, with a draconian nature of nationwide lockdown appears to be done relatively worse in preventing the number of confirmed cases compared to other countries with less restrictive regimes (3).

Nearly more than half of patients who have died of the infection have been aged 60 and above, and many have underlying conditions, hewing to the international data about older people being more vulnerable to the disease (4). At the same time, nearly, 13 percent of the Kerala population are aged over 60 (5). As per Economic Review by State Planning Board, around 27 percent men and 19 percent women are diabetic, 41 percent men and 39 percent women are hypertensive, more than one lakh people are being treated for cancer, 5.3 lakh people have chronic pulmonary disease and 4.8 lakh have Asthma (6). This paper compares the situation of COVID-19 pandemic in above mentioned major states and Kerala by the different phase of lockdown and also highlights Kerala’s fight against the pandemic.

### Kerala and the Pandemic

Kerala a small state in India, The land that proclaims itself as “God’s Own Country” is a heaven for tourists who make a beeline to the backwaters, palm-fringed sunset beaches, lush hills of the Western Ghats, and the spice plantations on the mountain slopes. Kerala has been known around the globe as a model for preventing the spread of COVID-19. Global media have been pouring praise on the state’s success (7,8). Kerala reported the first three cases of coronavirus in India in late January, all three victims being Indians who had studied in Wuhan. Later own Kerala received people from Italy too with positive cases including a whole family tested positive. The state soon began implementing mandatory quarantines for visitors arriving from abroad and from outside of the state, weeks before the Centre instituted similar measures across the country. A detailed guideline to deal with a new Coronavirus was issued by the Department of Health and Family Welfare of Kerala on 5th January 2020, much earlier than the first case detection in India. Importation and transmission-based approach for Testing Strategy On January 26th 2020, even before the first case of COVID-19 was reported in the state (9).

The critical situation is that Kerala is a small state (38,863 Sq.KM), but it homes for 35 million people makes it 819 people per square kilometer, eight most densely state in India. Other than people living in closely Kerala has 25 lakh migrants, who frequently travel to their native land. In addition to that, international travelling is a part of Kerala culture, which is connected to the rest of the world through four airports serve around 17 million passengers annually. Finally, Kerala has 2.5 million migrant laborers from other states. Simply Population density, affluent non-resident Keralites and thriving tourism all raise the risk for an outbreak in Kerala. However, the containment strategies adopted by Kerala have helped in slowing the spread of infection into the community by the end of May (10). Study also suggests that the head start that Kerala had in preventing the spread of the disease in the initial phase may help the state in dealing with the next wave of the disease (11).

Kerala opted for a combination of massive testing, quarantine and support of the poor with positive short-term effects (12). By January itself, the government started promoting using the face masks. On February 3, the state government declared Covid-19 as state calamity and drafted measures to be followed. By the beginning of March, government introduced more precautionary measures by shutting down the schools and colleges and asking all religious and other groups cancel all gatherings, including marriages, to encourage social distancing (13). On the nutritional point of view, free meals delivered for school children at their home. The government also instructed internet service providers to improve the bandwidth to encourage work from home (14). The use of traditional medicines for immune-boosting was encouraged, as Kerala is the land of Ayurveda.

#### Data and methods

We used publicly available data from https://www.covid19india.org/ and Covid-19 Daily Bulletin (Jan 31-May 31), Directorate of Health Services, Kerala (https://dashboard.kerala.gov.in/). Data compiled in this web portal is based on state bulletins and official handles. The details of the data are available on the website. This portal data matches with the data provided by the Ministry of Health and Family Welfare, Government of India (https://www.mohfw.gov.in/). For the analysis purpose, the number of Covid-19 cases categorized into 5 phases i.e., pre-lockdown (before 24^th^ March 2020), 1^st^ phase of lockdown (25^th^March to 14^th^ April 2020), 2^nd^ phase of lockdown (15^th^ April to 3^rd^ May 2020,) 3^rd^ phase of lockdown (4^th^ May to 17^th^ May 2020), and 4^th^ phase of lockdown (18^th^ May to 31^st^ May 2020).

We calculate the phase-wise period prevalence rate (PPR) and the case fatality rate (CFR) of the last phase. The measures are,

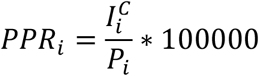

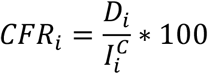

Where 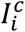 the total Covid-19 is confirmed cases in the i^th^ lockdown phase; Pi is the total population of the state and Di is the number of deaths i^th^ lockdown phase. It must be noted that a robust estimate of CFR is possible only at the end of the pandemic. However, the dynamics of fatality and recovery rates across affected countries and states would enhance the knowledge base and provide useful information in the ongoing outbreak of Covid-19.

## Results

The pandemic is among the worst to have happened in the history of mankind. The disease a little later started sweeping across all the Indian states adding up to a dreadful toll. Change in cumulative caseload over time in India (Figure 1) shows that the infections increased rapidly since March and there was a sharp rise of cases in May 2020. On the other hand, for a population of over a billion, India has approximately 1.9 million hospital beds, 95 thousand ICU beds, and 48 thousand ventilators as estimated by a research team (15). The study reveals that most of the beds and ventilators in India are concentrated in seven states; Uttar Pradesh (14.8%), Karnataka (13.8%), Maharashtra (12.2%), Tamil Nadu (8.1%), West Bengal (5.9%), Telangana (5.2%) and Kerala (5.2%).

**Figure 1:**
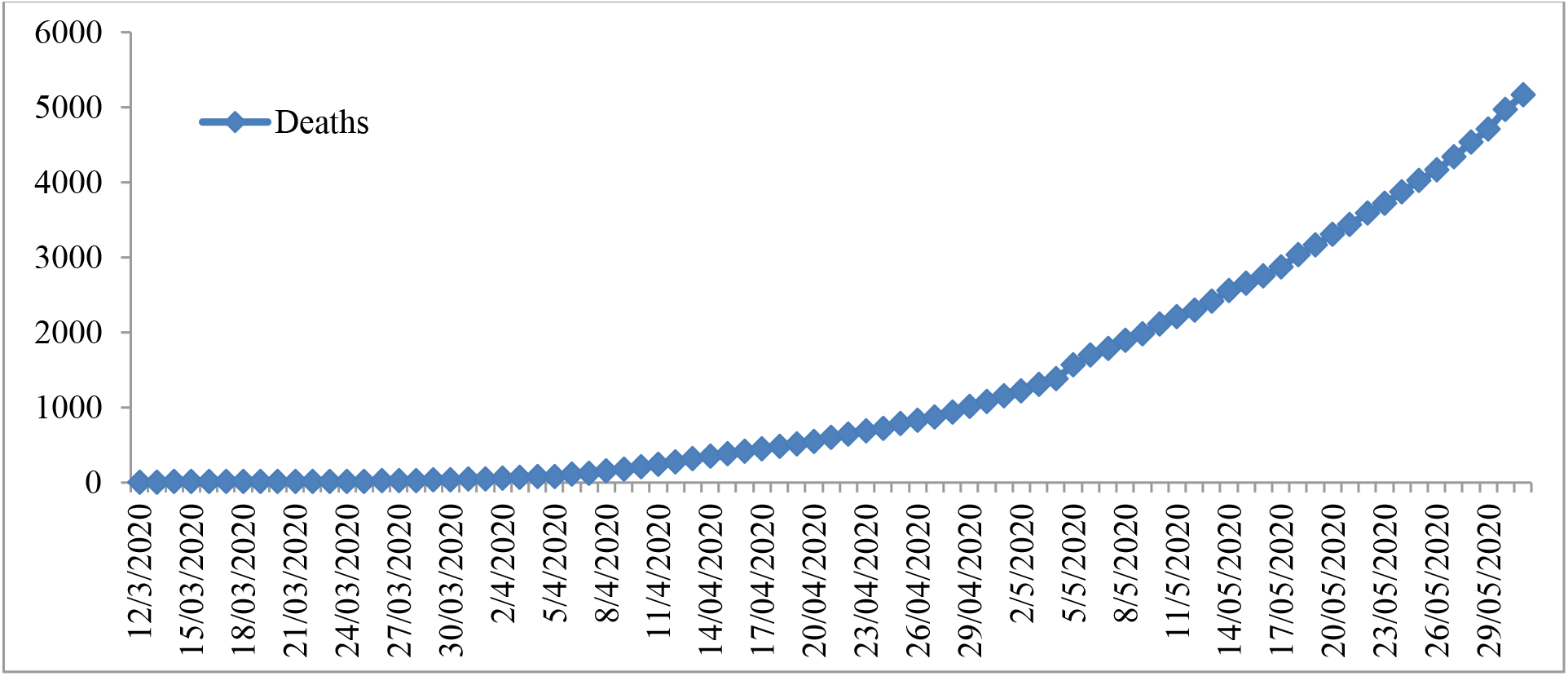
Distribution of cumulative caseload over time. **Source: https://covidindia.org/**

State-wise distribution of cases in India (Figure 2) reveals Maharashtra to have the highest burden of cases with highest number of confirmed and death cases followed by Tamilnadu, Delhi and Gujarat. Whereas, in Kerala, where the epidemic started early in the country, the number of confirmed and death cases were the least.

**Figure 2:**
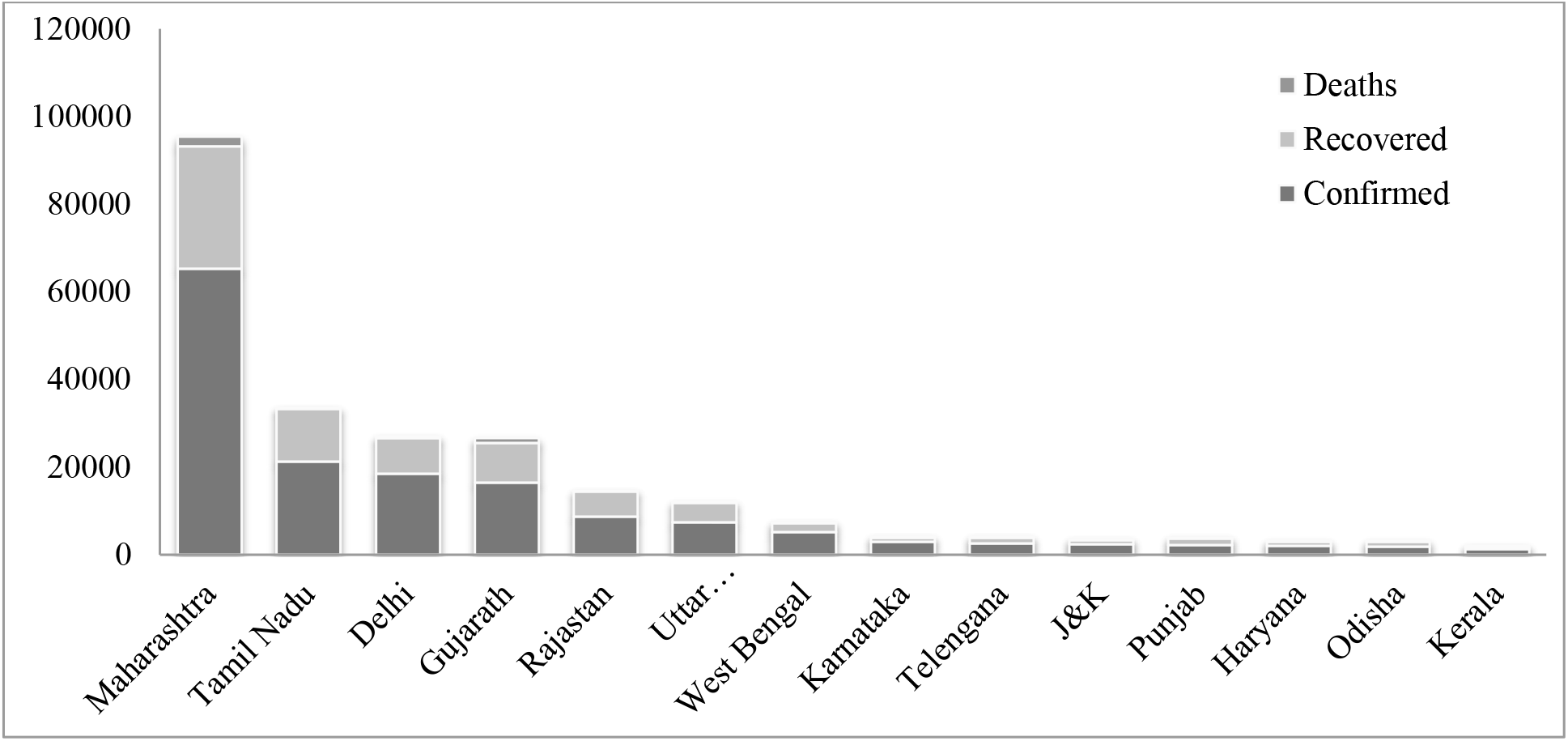
State-wise distribution of Covid-19 confirmed, recovered and death cases. **casesSource: https://covidindia.org/**

Figure 3 presents the pattern of new cases of COVID-19 in period prevalence rate in major states. The maximum new cases were found in Maharashtra until the end of the fourth lockdown period. The total number of new cases has increased consistently nearly in all states over the lockdown period. However, Kerala is the only state experiencing a decline in new cases per day till the third lockdown. Then, Kerala also experienced a spike in the fourth lockdown in the month of May. Results also show that among the most affected states, Maharashtra and West Bengal stand at the top with higher levels of CFR as against Kerala. It is also found that Kerala recorded negative percentage change in the new COVID-19 cases between lockdown-1.0 and lockdown-2.0 and between lockdown-2.0 and lockdown-3.0 (16).

**Figure 3:**
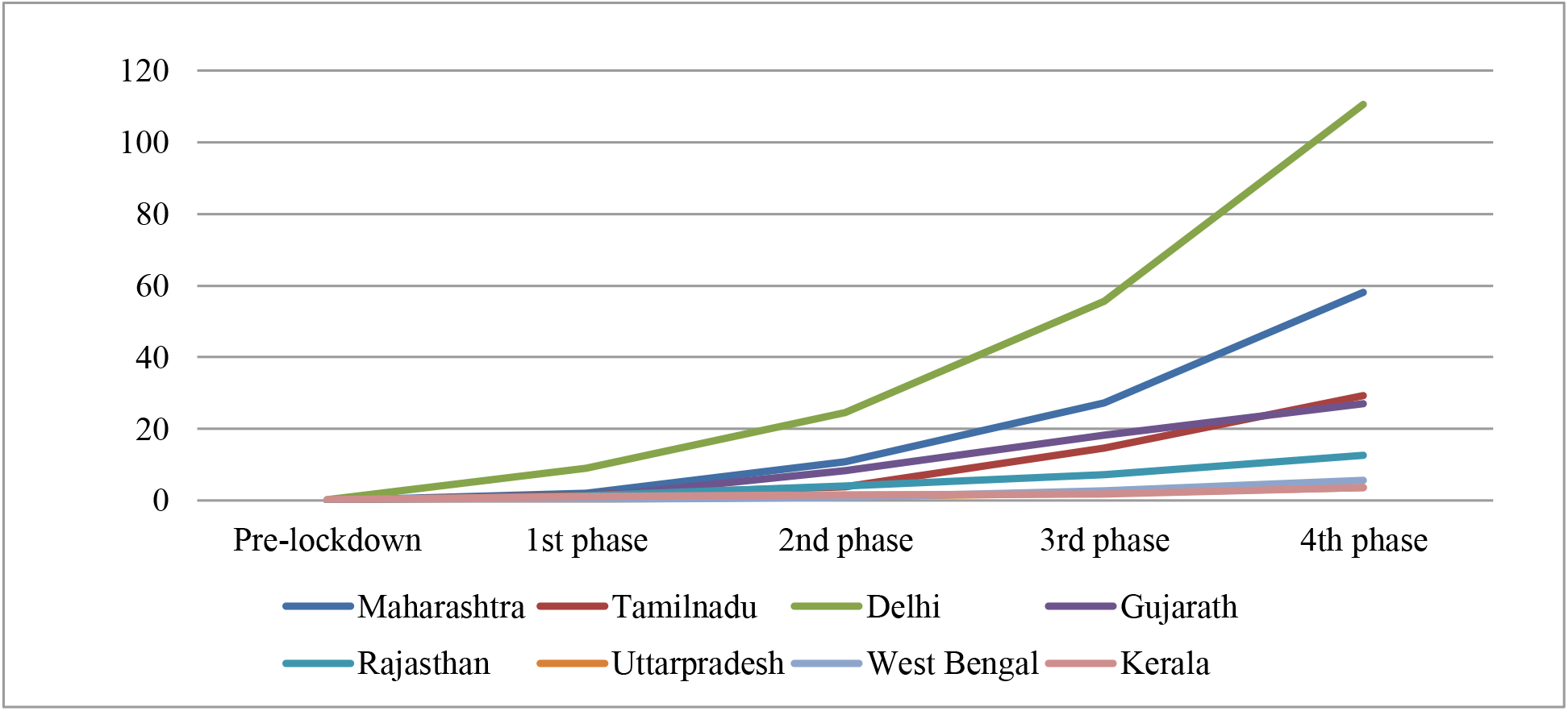
Period prevalence rates of major states hit by Covid-19 by different lockdown phases. **Source: https://covidindia.org/**

**Figure 4:**
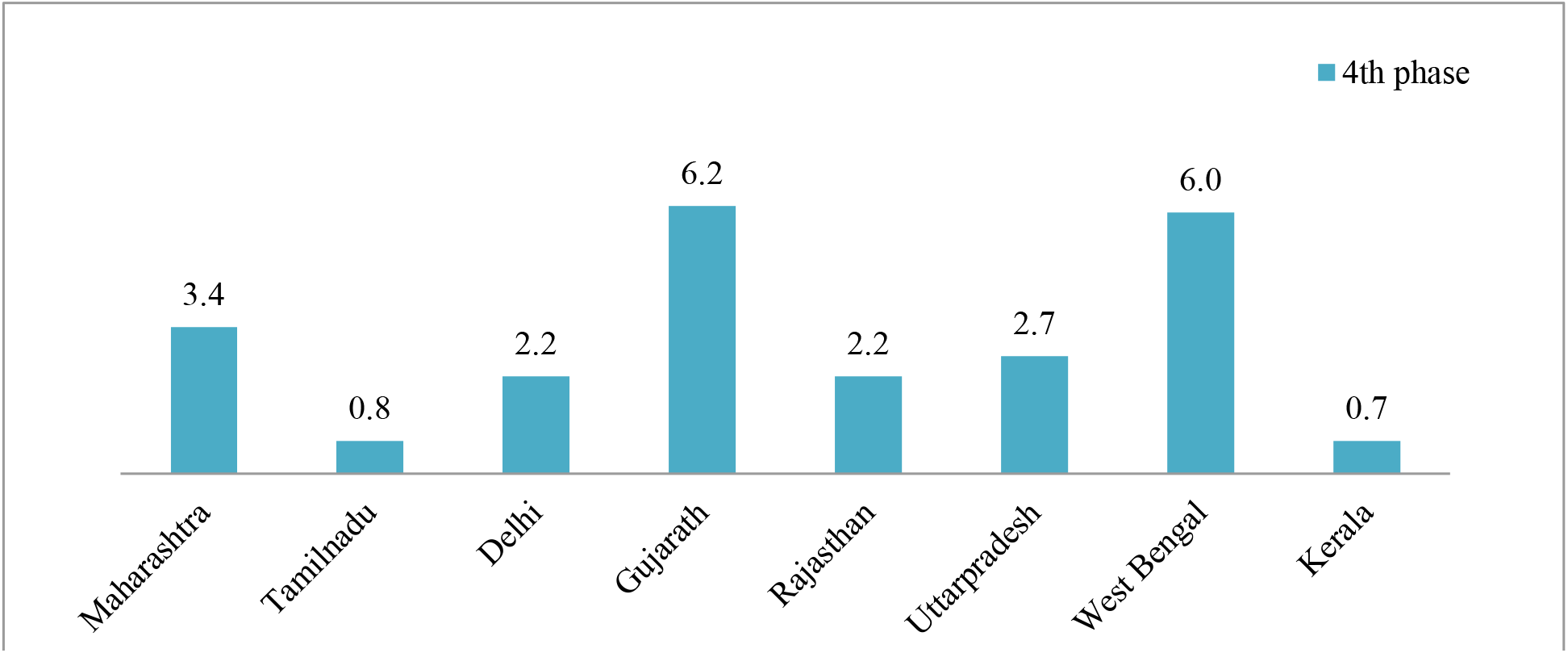
Case fatality rates of major states hit by Covid-19 in the last lockdown phase. **Source: https://covidindia.org/**

The state of Kerala, where the epidemic started early in the country, has only less than one percent of case fatality rate. While among other most affected states, fatality rate is 6.2 percent in Gujarat and 6 percent in West Bengal.

### Kerala: COVID-19 response

Kerala has been known around the globe as a model for preventing the spread of COVID-19. Global media have been pouring praise on the state’s success. The state reported the first case of the country in January. The state moved fast because of that by mid-January it had already put in place a strategy to isolate people who showed symptoms in hospitals, to trace their contacts and put them in-home quarantine. Kerala has done remarkably well in controlling the virus transmission, even compared with countries with similar populations (Table 1). Also compared to countries like Poland (27,365), Canada (97,779) and Netherlands (47,945), Kerala reported only 1261 Covid-19 cases by the end of May 31. Malaysia populated the same as Kerala, known for its strict and complete successful shutdown, reported seven times more number of cases compared to Kerala.

**Table 1.** Population and Covid-19 cases of Selected Countries and Kerala.

| Country | Poland | Iraq | Canada | **Kerala** | Peru | Morocco | Malaysia | Saudi Arabia | Uzbekistan | Netherlands |
| --- | --- | --- | --- | --- | --- | --- | --- | --- | --- | --- |
| Population | 37.1 | 36 | 35.5 | **35** | 33.3 | 33.3 | 33.2 | 30.7 | 30.4 | 17.8 |
| Covid-19 Cases (May 31) | 27,365 | 13,481 | 97,779 | **1261** | 1,99,696 | 8,408 | 8,329 | 105,283 | 4,448 | 47,945 |

Source: John Hopkins University, Kerala Government and UN Population.

Four months on, Kerala is being praised for not just flattening the curve of the deadly infection even as it shoots up in many parts of the country but for having an extremely low mortality rate and high recovery rate. What is the strategy behind the success story?

#### Testing

Identifying the case from the beginning got special attention; intensive training provided to health workers varies from doctors to Asha workers to identify the Covid-19 symptoms. Health check-up booths established in all railway stations, state borders and airport the check and guide the people coming from outside. For speedier testing, the government set up a facility at virology institute Alappuzha with the help of National Institute of Virology, Pune. Soon after 12 more testing labs were launched in different parts of the state. In addition to using the centrally procured real-time polymerase chain reaction (PCR) testing kits, Kerala was the first state to acquire rapid test kits from the Pune-based Mylab.

The state used testing optimally, figure 5 shows the number of COVID-19 tests and active cases in a different phase of lockdown in Kerala, The number of tests and active cases are showing non-linear trend throughout the period but in Phase 4 of lockdown both showing an increasing trend. The number samples tested per day never crossed 1000 till the beginning of the May only four times more than 1000 tests conducted, the average number of tests per day after 100 active cases were 687 till the end of Phase 2 (Fig 5). One active case per 84 tests was also reported till May 31. In phase 3 number, tests increased to more than 1000 per day due to the return of the people from different part of the country and the world. The average number of tests increased from 721 in the first two phases to 1214 in the last periods of the lockdown. Till the end of the lockdown, the average number of tests per day stood below 600. With the rise of active cases, number of tests per day also increased.

**Figure 5:**
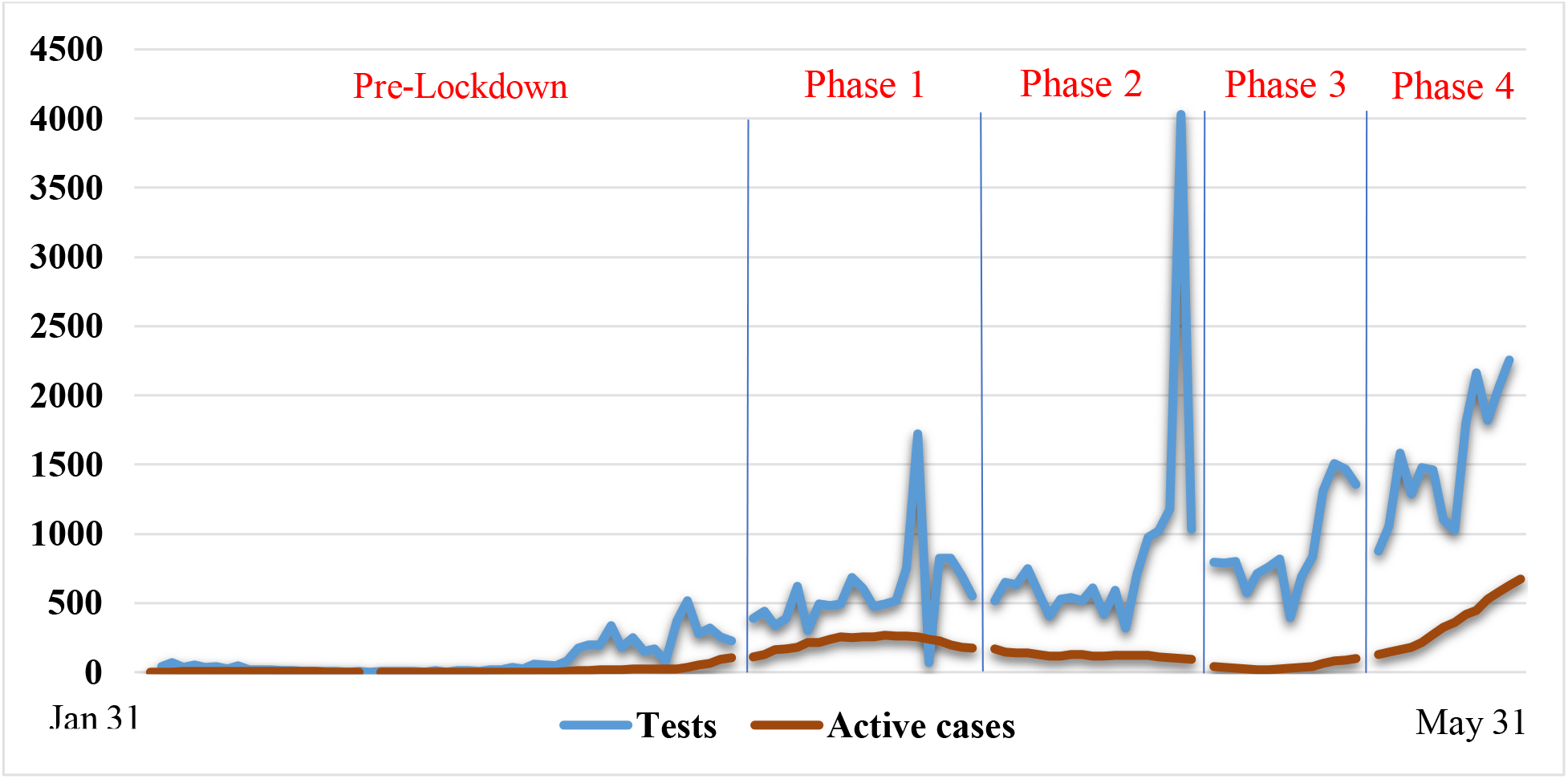
Number of Covid-19 Tests and Active Cases in Different Phase of Lockdown-Kerala. **Source: Covid-19 Daily Bulletin (Jan 31-May 31), Directorate of Health Services, Kerala.**

Tests are conducted optimally with prioritizing the groups to be tested rather than the whole population. This selective approach was taken due to the Limiting factors include availability, reagents, PCR equipment, trained staffs, protective gear for each collection and cost (Rs 4,500 per test). The Indian Council of Medical Research (ICMR) handed out 100,000 RDKs to Kerala in the beginning; Instead of testing the general population, the Kerala government identified four priority groups. 1) Healthcare workers who had served patients with COVID-19 and other patients (25,000 kits). 2. Government staff with public contact, like police personnel, ASHAs, Anganwadi workers and local government staff (20,000 kits), and Essential service providers (5,000Kits), e.g., -workers of community kitchens, food and grocery deliverers, and ration shop vendors, etc. 3) People quarantined at home (25,000 kits). 4) All senior citizens (20,000 kits). At the beginning (first phase lockdown) one active case per 34 tests reported (Fig 5) due to the formidable step was taken by the government.

#### Tracing

Contact tracing was conducted effectively by devolved power and funding to grass root-level bodies, and established a social system that promotes community participation and public cooperation. Other than that, 2.5 lakh volunteers (Age group 22-40) were raised through government registration and trained with local government bodies and deployed to deliver the food, contact tracing and checking those under in quarantine. People who had returned from infected countries after January 14 were tracked and scanned by the help of local government bodies. By the beginning of the march introduced the mandatory quarantine for 28 days for everyone who enters the state and more helplines were created at local levels including a separate helpline for mental health. High risk people who were in contact with the positive cases were tracked, and kept in quarantine helped in optimal utilization of the Covid-19 treatment kits. For the 300 positive cases reported in Kerala in April, close to 1.75 lakh people were traced and quarantined.

Figure 6 shows the number of people isolated per one active COVID-19 case in a different phase of lockdown in Kerala. Dynamic tracing will help the government to utilize the available recourse to fight the pandemic. Throughout the time, an average number of 581 people kept in isolation per active case. It varied in a different phase of the lockdown, it was high in the beginning, to avoid the risk of community spread. Then it reduced to fewer numbers in first two phases of lockdown due to the uncompromising regulations to prevent the social contact while in third phase isolation per active case got increased due to lower number of active cases in that state (Fig 6). In the fourth phase as the number of active cases increased the isolation per active case was also reduced, the compulsory isolation for whoever entering the state in this period reduced the risk social contact and the high number of isolation.

**Figure 6:**
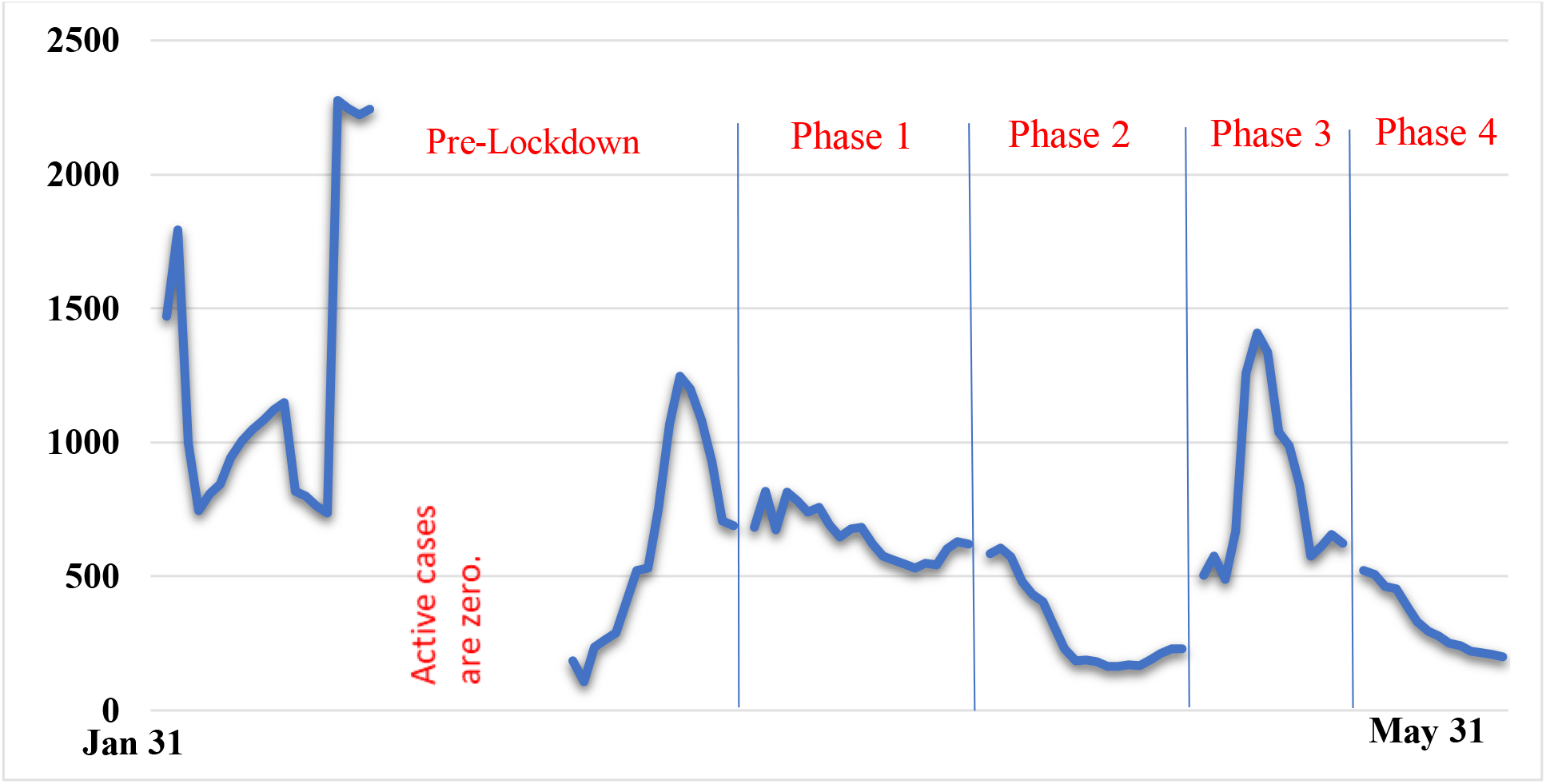
Number of People Isolated per One Active Covid-19 case in Different Phase of Lockdown-Kerala. **Source: Covid-19 Daily Bulletin (Jan 31-May 31), Directorate of Health Services, Kerala.**

Government set up isolation wards in all medical colleges, district and general hospitals besides around 650 corona care centers were created in hostels, educational institutions and unoccupied buildings. Around 1lakh room were kept aside to face emergencies. The district Kasaragod was the highest number of Covid-19 cases registered in Kerala, the government opened medical college hospital in two days with more than 250 beds, well-equipped machinery and staffs. By joining with Tata group, the government of Kerala is developing a 500-bed intensive Covid-19 treatment center at the same district.

Figure 7 shows the percentage of people in home isolation in different phases of lockdown in Kerala. It points that 9 out of ten people were isolated in their own home with necessary support including food, medical check-up and counselling and the government was ready with enough number of isolation beds to accommodate a large number of people. The people who were not able to self-quarantine themselves their own house, the government didn’t directly admit them in the hospital facilities; they have been admitted in the isolation facilities created in hostels, educational institutions and unoccupied buildings. Those who showed symptoms were admitted to hospital facilities and got tested.

**Figure 7:**
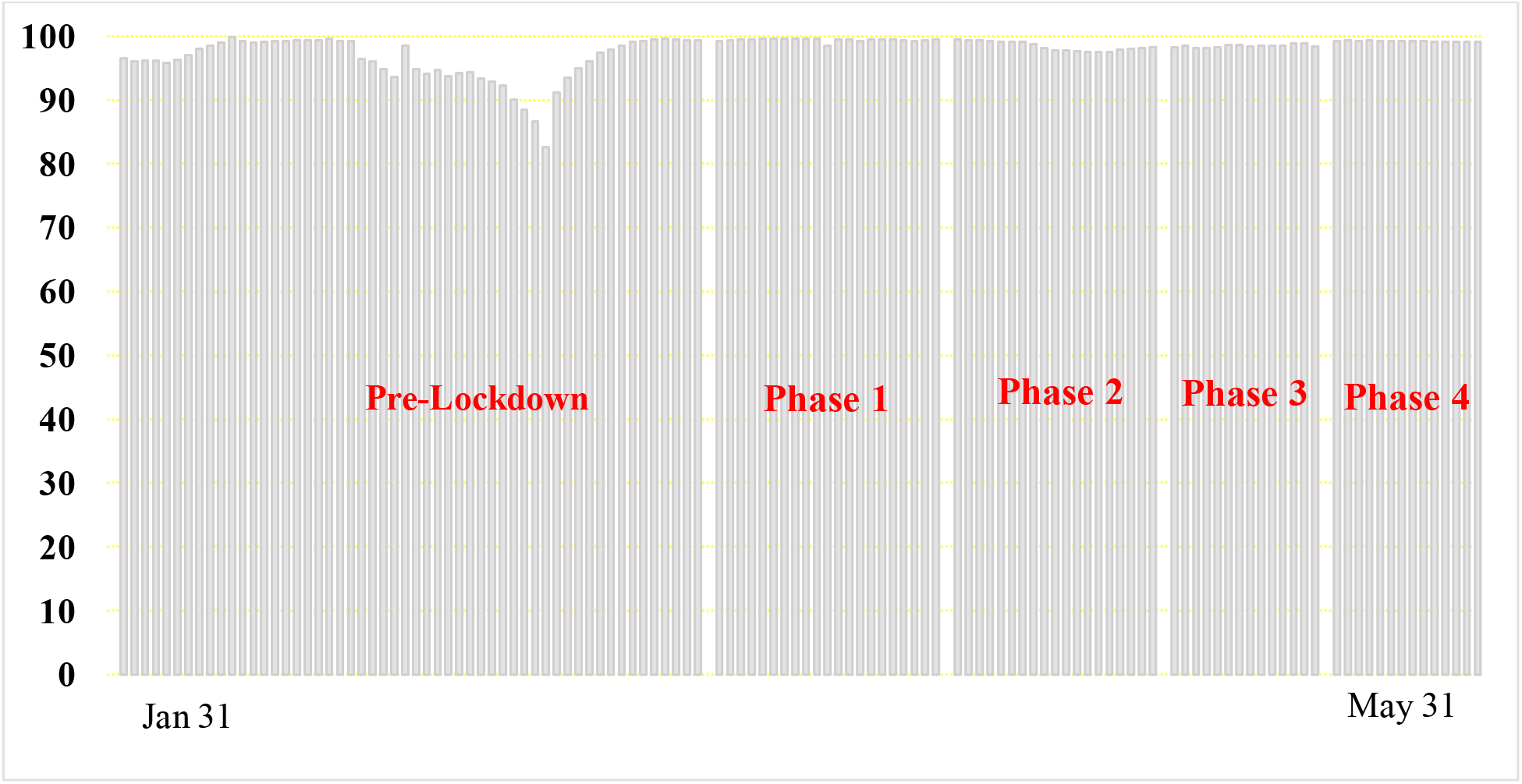
Percentage of People in home isolation in Different Phase of Lockdown-Kerala. **Source: Covid-19 Daily Bulletin (Jan 31-May 31), Directorate of Health Services, Kerala.**

#### How this systematic testing and tracing helped the Kerala government

Figure 8 shows the number of hospitalization related COVID-19 in different phases of lockdown in Kerala. This intensive approach (Systematic contact tracing and isolating) help the health system from being overburdened by people, In average 76 people hospitalized per-day because of that at its peak, only 816 people (In April-Phase 1) and 1241 people in the fourth phase were admitted. Average of 96 people per day hospitalized in all phases, but in the last phase, it got increased to 181 people per day due to the return of the people from outside of the state (Fig 8). Even at the end of May per day, hospitalization never crossed 250 marks and had less than 1500 people admitted to health facilities all the time.

**Figure 8:**
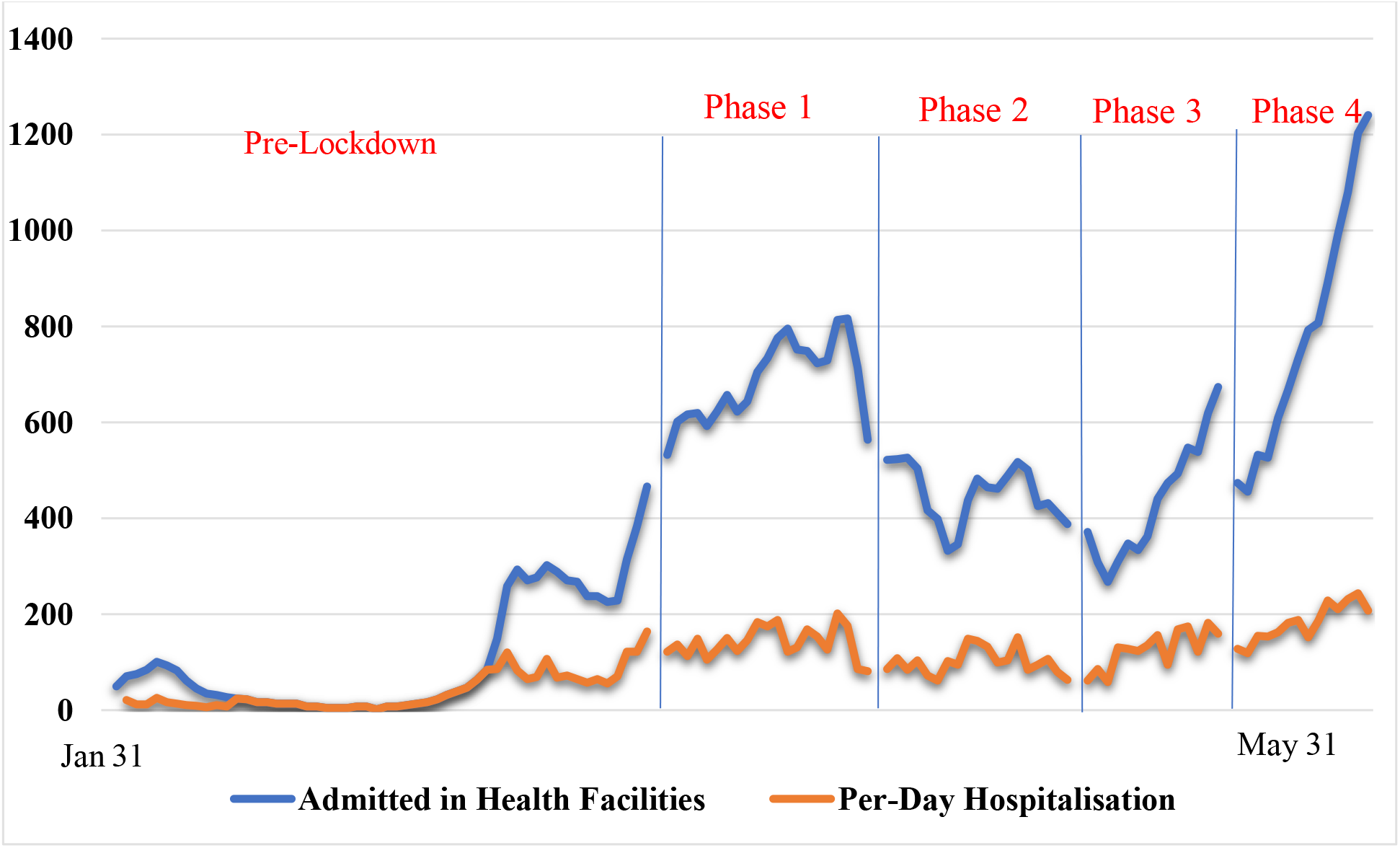
Number of hospitalizations related Covid-19 in Different Phase of Lockdown-Kerala. **Source: Covid-19 Daily Bulletin (Jan 31-May 31), Directorate of Health Services, Kerala.**

### Social Distancing and Other Measures

March 23, Kerala announced complete lockdown before the announcement of national lockdown. The most crucial step was taken on March 15 by announcing an awareness campaign named ‘Break the Chain’ to create awareness and promote social distancing. The government was holding a press conference every evening to answer all the queries related to the outbreak and to reduce the spread of rumors. Local bodies mandate that everyone hold an umbrella when they go out, and distributed umbrellas produced by Kudumbashree to promote involuntary social distancing. In high-risk areas, the police force created an online delivery system of essential food items. Innovative products have been developed as part of beating the Covid-19 in the state. A smart bin called ‘BIN-19’ has been launched for the collection and disinfection of Used Face-Mask which is based on Internet of Things (IoT) (17).

Kerala became the first state that does away with ‘zonal classification of districts on the basis of Covid-19 spread where more police would be deployed to ensure the strict adherence of quarantine and lockdown norms (18). Meanwhile, Kudumbashree, a Keralan grassroots network of local organization and women’s self-help groups, has helped the state’s containment strategy by producing masks and hand sanitizer. Some 1,200 community kitchens were established to feed the indigent and unemployed, and Kudumbashree has already served 300,000 meals a day. Kerala’s educated population has behaved responsibly, limiting community transmission, and cooperating with authorities.

One of the two major challenges that Kerala faced in this period was the security of the migrant workers from other states, just after the announcement of the national lockdown, the state set up nearly 20000 camps for migrant laborers, by calling them, ‘guest workers’. 3 lakh workers were hosted in these places and urged to wait in place till the special trains will be arranged to take them home. The second challenge was the returning migration from abroad; they were tested before taking the flight and rechecked after landing in Kerala. Once they reach Kerala, they will be taken to government facility for quarantine in respected districts and requested to stay there until the time of quarantine is over. Same time government set up isolation wards in all medical colleges, district and general hospitals besides around 650 corona care centers were created in hostels, educational institutions and unoccupied buildings.

### Conclusion

The state got into the action by January and rolled out measures that reduced the spread of the Covid-19. The first coronavirus case in India was confirmed in Kerala’s Trissur district on January 30, 2020. Ever since, the state has been fighting the pandemic in an exemplary manner. The steps were simple but proactive. The equation for Kerala’s success has been simple, prioritized testing, widespread contact tracing, and promoting social distance. Besides that two factors fuelled this success, the ‘formidable’ grass root level primary health care system and the experience it gleaned in the last two years when it handled another deadly virus outbreak (2018 Nipah virus outbreak in Kerala).

State announces its strategy is trace, quarantine, test, isolate and treat in the beginning only. The state soon began implementing mandatory quarantines for visitors arriving from abroad and from outside of the state, weeks before the Centre instituted similar measures across the country. They also imposed uncompromising controls, were supported by an excellent healthcare system, government accountability, transparency, public trust, civil rights and importantly the decentralized governance and strong grass-root level institutions. This much decentralized system has hold out the test of two severe floods and another viral outbreak in past years, ordinarily making good use of the active and voluntary engagement of the public.

While the rest of India worries about the rate at which new cases are doubling, that question has become almost irrelevant in Kerala, the curve has been flattened for now and transmission limited. Currently, how it manages to ease the lockdown safely will depend on a large number of factors. The summer monsoon rains, the floods that will follow, as well as returning migrants will add layers of complexity. But one thing is clear: when the next wave of novel coronavirus hits, which it will, the state will be ready. The “proactive” measures taken by Kerala such as early detection of cases and extensive social support measures can be a “model for India and the world”

## Data Availability

All the data referred to in the manuscript are available in the public domain

https://www.covid19india.org/

https://dashboard.kerala.gov.in/

